# Direct targeting for focused ultrasound thalamotomy in the treatment of movement disorders: a retrospective cohort study

**DOI:** 10.1101/2025.09.03.25335065

**Authors:** James Cahill, Nemanja Useinovic, Andrew Toader, Hanna E. Minns, Matthew Henn, Adam Lipson, Daniel R. Cleary

## Abstract

**Background:** Accurate targeting of the ventral intermediate nucleus (Vim) remains a critical challenge in stereotactic thalamotomy for essential tremor (ET) and tremor-dominant Parkinson’s disease (TDPD). Indirect atlas-based methods suffer from interindividual anatomical variability and poor visualization of thalamic substructures. We evaluated the clinical impact of a direct targeting strategy enabled by fast gray matter acquisition T1 inversion recovery (FGATIR) imaging in MR-guided focused ultrasound (MRgFUS) thalamotomy.

**Methods:** We conducted a retrospective cohort study of adult patients who underwent first-time MRgFUS thalamotomy for ET or TDPD at the Oregon Health and Sciences University (Portland, Oregon) between August 2023 and January 2024. Patients treated with FGATIR-guided direct targeting (n=64, male=40) were matched to a cohort treated using indirect targeting combined with physiological mapping (n=52, male=36). Data was collected from intraoperative recordings and postprocedural imaging, as well as one-day, one-month, and three-month follow-up evaluations. Procedural efficiency, lesion and edema characteristics, tract involvement, clinical outcomes, and adverse events were assessed.

**Findings:** FGATIR-guided direct targeting significantly reduced the number of sonications (−22%), total sonication time (−32%), and overall procedural duration (−33%) compared to indirect targeting. Lesion volumes and perilesional edema were smaller in the direct targeting group with less impingement on the internal capsule and medial lemniscus. Direct targeting had higher initial sonication accuracy with a lower incidence of neurologic deficits. Both groups achieved similar tremor improvements.

**Interpretation:** FGATIR-based direct targeting improves the safety, precision, and efficiency of MRgFUS thalamotomy without compromising clinical benefit. The FGATIR sequence is widely available on clinical MRI systems, and this method may be adapted for use with other targets for stereotactic ablation in functional neurosurgery. Direct targeting represents a scalable and patient-centered advancement for stereotactic thalamotomy with minimal technological barriers to widespread adoption.

**Funding:** OHSU Parkinson Center Pilot Program; Oregon Medical Research Foundation #1029114.

## Introduction

Essential tremor (ET) and tremor-dominant Parkinson’s disease (TDPD) are highly prevalent movement disorders that are disabling in advanced stages. The development of MR-guided focused ultrasound (MRgFUS) has driven a resurgent interest in thalamotomies as a safe and effective treatment for movement disorders.^1,2^ MRgFUS is an incisionless technique that provides immediate and durable tremor control comparable to stereotactic radiosurgery, gamma knife, and deep brain stimulation (DBS) but with fewer complications and without the need for anesthesia or follow-up programming.^3-5^ One persistent limitation of all these procedures has been optimal targeting of the ventrointermediate nucleus (Vim).^6^ Accurate targeting of the Vim is crucial, as it measures only 4 mm by 4 mm by 6 mm and is bordered by neurologically important structures,^7^ damage to which can result in weakness, sensory changes, and imbalance. The standard targeting method uses atlas-derived coordinates to indirectly approximate the location of the Vim, followed by iterative trial-and-error physiological mapping. This approach is limited by the rapidly increasing energy requirement of successive sonications,^8,9^ which increases risk of complications or incomplete treatment.^10,11^

To address this limitation, advanced imaging techniques have been explored to directly identify the Vim. To date, however, direct targeting methods suffer from significant limitations, ranging from poor spatial resolution, imaging artifacts, and inconsistent results, to expensive equipment requirements and computationally intensive data processing.^7,12-17^ In contrast to prior methods, the fast gray matter acquisition T1 inversion recovery (FGATIR) sequence is a conventional and readily available MRI sequence capable of producing high fidelity anatomical images through enhanced white-gray matter distinction.^18^ FGATIR can be leveraged to identify a direct target for thalamotomy within the dentatorubrothalamic tract (DRTT) fibers, where tremor suppression is maximized while limiting collateral involvement.^7^ This target can be visualized on FGATIR as the interface of the superolateral aspect of the rubral wing (RW; the FGATIR anatomical correlate of the DRTT) with the inferolateral surface of the thalamus.

## Methods

### Study design

This is a retrospective cohort study of MRgFUS Vim thalamotomy for tremor treatment, using direct versus indirect Vim targeting. All patients were treated at a single academic center in North America (Oregon Health & Science University, Portland, Oregon, USA) by a single attending neurosurgeon (DRC). Collected data included outcomes from all eligible patients treated between August 2023 and January 2025. The study protocol was reviewed and approved by OHSU’s Institutional Review Board (STUDY00026905).

### Patients

Patients undergoing MRgFUS thalamotomy treatment for tremors at OHSU during the two-year period were identified from electronic medical records. Patients were aged 18 years or older, had been diagnosed with ET or TDPD by a neurologist or neurosurgeon, had disabling upper extremity tremor, and were medically refractory to at least two pharmacologic therapies. Patients who had previously undergone ablative treatment of the basal ganglia for any reason were excluded from the study. Exclusion criteria were kept minimal to reduce selection bias. All patients provided written informed consent prior to their procedure.

### Procedures

All demographic and clinical history, including sex and ethnicity attributes, was sourced from medical records. Outcome data was sourced from pre- and postoperative tremor evaluation at time of procedure, postoperative day one (POD1) imaging, and POD1, one-month, and three-month postoperative assessments.

From August 2023 to June 2024, MRgFUS thalamotomy was performed in standard fashion.^19^ Prior to treatment, the patient’s scalp was shaved, a stereotactic head frame was fixed to the head under local anesthesia, and a plastic diaphragm was fitted just superior to the brow line. The patient was laid supine in a 3T MRI scanner with a 1024-element phased-array ultrasound transducer attached to the head frame and plastic diaphragm. T2 and FGATIR sequences were acquired. Treatment planning was done using Exablate software (InSightec, Haifa, Israel), and treatment temperatures were measured using MR thermometry.

For the indirect targeting cohort (n = 52), initial target localization was accomplished using standard stereotactic coordinates: 28% of the AC-PC line anterior to the PC, 11 mm lateral to the third ventricle, and 1·5 mm superior to the AC-PC plane. Following alignment, low-energy sonications to 45-49 C were used to test for tremor improvement and side effects, including weakness, ataxia, dysarthria, and sensory disturbances. Adjustments were made to the target location, and the process repeated until sufficient tremor control was achieved with no or minimal side effects. The permanent lesion was created through a target temperature of 55-56 C for 1-2 seconds, or a lower temperature for longer duration.

Patients in the direct targeting cohort (n = 64) were treated between July 2024 and January 2025. Instead of standard indirect coordinates, the initial treatment target was placed directly at the junction of the superolateral aspect of the RW with the inferolateral aspect of the thalamus (Figure 1B). The target was adjusted to be 3·5 mm from the medial boundary of the internal capsule (IC) in the axial view, 4 mm from the curvature of the IC in the coronal view, and 4 mm from the anterior border of a white matter tract aligned to the medial lemniscus (ML) in the sagittal view (Figure 1A). Verification sonications, physiological testing, and permanent lesioning proceeded as per indirect targeting.

**Figure 1.**
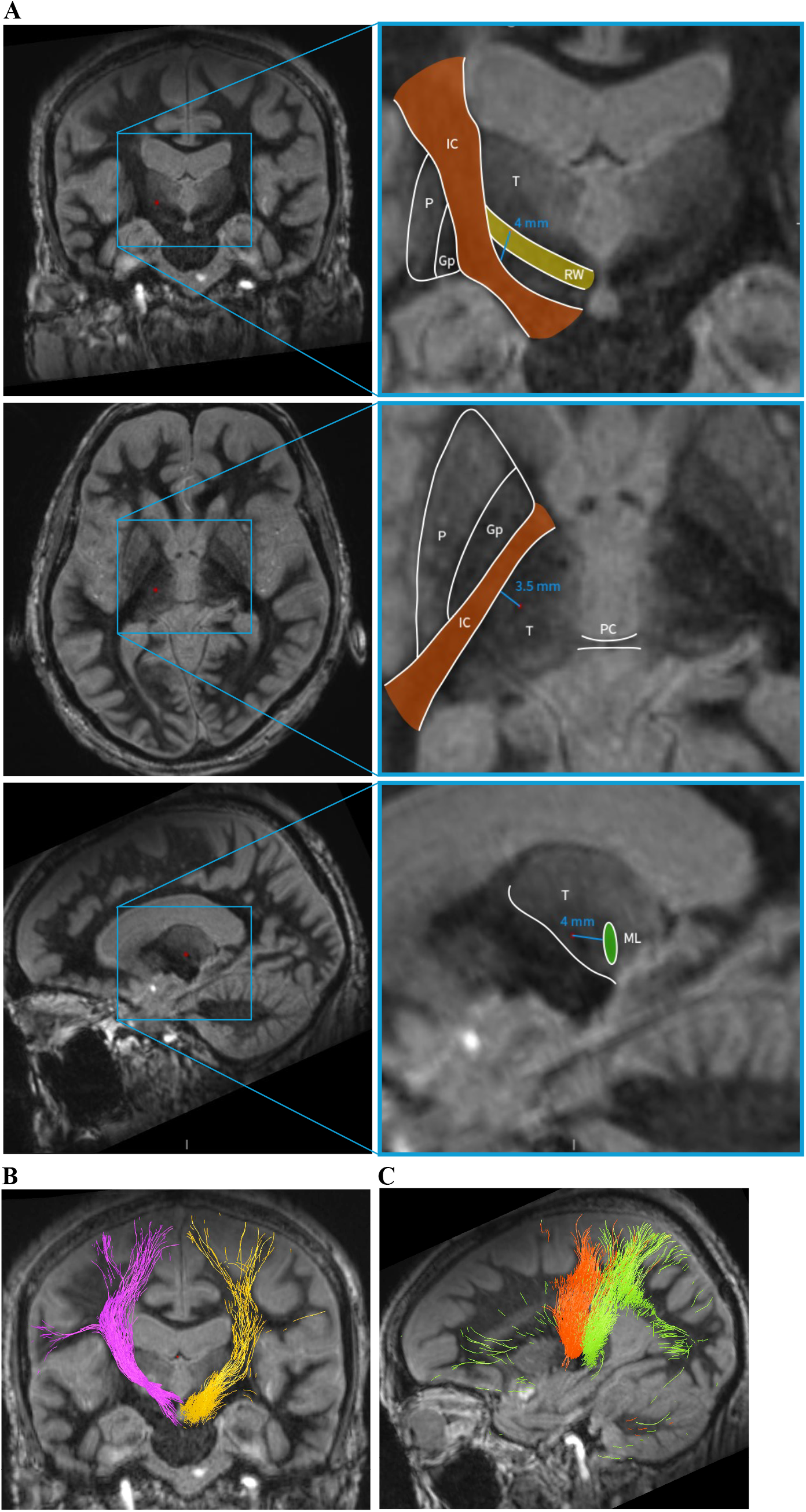
FGATIR-enabled direct targeting. (A) FGATIR sequences of coronal, axial, and sagittal slices orienting target location (red dot) within the greater cerebral anatomy (left column), and relative to surrounding white matter structures used as reference points to further constrain targeting parameters (right column). (B) Diffusion tensor tractography isolated fibers seeded at the target location (purple) are compared to dentatorubrothalamic (DRTT) fibers seeded at the contralateral cerebellar dentate nucleus (gold), supporting the identification of the ablated fibers as part of the DRTT. Fibers were overlaid onto a post-operative day one FGATIR sequence, allowing for visualization of the lesion. (C) Seeds in the internal capsule (corticospinal tract, orange) and a white-matter structure putatively identified as the medial lemniscus resulted in tracts running to the primary motor and primary sensory cortices respectively, in keeping with their established physiologic function. Tracts in B and C are color-matched to their respective anatomical correlates in the right-side column of A. T = thalamus; P = putamen; GP = globus pallidus; IC = internal capsule; PC = posterior commissure; ML= medial lemniscus.

### Outcomes

Procedural parameters, including sonication time and energy imparted, were recorded in the InSightec Exablate software and exported for analysis.

Radiological assessments of thalamotomy-induced lesion and perilesional edema were quantified using POD1 imaging. Vector distances of the lesion center in the anteroposterior, mediolateral, and superoinferior directions relative to the posterior commissure (PC) were taken. Lesion size was measured on T1 sequences, perilesional edema was measured on FGATIR sequences (Supplementary Figure 1), and width of non-impinged ML was measured on FGATIR sequences. IC and RW impingements were scored according to a predefined 6-point ordinal scale, and ML impingement was scored according to a 3-point ordinal scale (Supplementary Figure 2).

For visualization of individual tracts, diffusion tensor tractography was used to isolate pathways of interest (Stealth Medtronic Tractography, Medtronic, Inc) which were then overlaid onto FGATIR sequences (Figure 1B,C). Tractography seeds were placed at the target coordinates, as well as within the dentate nucleus, ML, and IC. Tracts seeded at the target coordinates and in the dentate nucleus of the cerebellum showed strong overlap with the RW on FGATIR. (Figure 1B). Tracts seeded in the adjacent IC spread to both the motor cortex and supplementary motor area, while tracts seeded in the ML extended to the sensory cortex (Figure 1C).

Clinical evaluation of baseline tremor severity was performed through patient history, written assessment tools, and focused neurological examination. Intraoperative assessment of tremor improvement consisted of spiral drawing tasks and clinical evaluation of speech, strength, and sensation. Post-operative assessment included spiral drawing tasks and focused neurologic examination. Clinical notes from POD1, one month, and three months follow-ups were reviewed for patient-reported adverse effects and subjective tremor control, which was scored post-hoc using a 3-point ordinal scale: significant improvement, moderate improvement, and no improvement.

Quantitative assessment of written pre- and postoperative spiral drawings was performed using custom MATLAB (MathWorks, Natick, Massachusetts, USA) pipeline, described in detail previously.^20^ Briefly, scanned spirals were segmented and separated from the background template. The spiral path was unraveled into a one□dimensional profile, and oscillations in spatial frequency-domain were extracted. Spatial power spectral density (PSD) was calculated using Welch’s method. A quantitative tremor score was derived from the number of intersections between the drawing and the spiral path, the length fraction (ratio of spiral length to the template length), and area under the curve for PSDs. Percent improvement is based on relative change in tremor score between pre- and post-operative drawings.

### Statistical Analysis

All statistical analyses were performed in GraphPad Prism v.10.6.0. Group comparisons for continuous variables were made using unpaired *t*-tests with Welch’s correction and Mann–Whitney *U* tests. Categorical and ordinal outcomes were compared using Fisher’s exact tests and Cochran–Armitage trend tests. Continuous variables were evaluated using Spearman rank correlation coefficients (ρ). Linear regression was used to model directional relationships and predictive strength (*R^2^*). Model fits were evaluated using *R^2^*, residual standard deviation (Sy.x), and visual inspection of residual plots. Runs tests were used to assess deviation from linearity. Logistic regression models were used to assess binary outcomes (e.g., presence of neurologic deficits), with model performance reported as odds ratios (ORs), 95% confidence intervals, and areas under the curve (AUC). Kruskal–Wallis tests and ordinal logistic regression were used to evaluate ordinal data. Data were reported as mean ± SD and graphed as mean ± SEM. Statistical significance threshold was p < 0·05.

### Role of Funding Source

The funders of the study had no role in study design, data collection, data analysis, data interpretation, or writing of the report.

## Results

### Cohort Characteristics

A total of 116 patients were included, with 52 patients treated using indirect targeting and 64 patients treated with direct targeting. Detailed patient metrics and clinical comorbidities are summarized and compared in Table 1. No statistically significant demographic differences or comorbidities were observed between the groups. The majority of patients underwent left-sided Vim thalamotomy for a right-handed tremor (indirect: 78·8%; direct: 71·9%). ET was the predominant diagnosis in both groups (indirect: 82·7%; direct: 78·1%), with the remainder diagnosed with TDPD. Preoperative skull density ratio (SDR) did not differ significantly between groups (Table 1).

**Table 1.** Demographic and clinical characteristics.

|  | GROUP |  |  |  |  |  | p-values |
| --- | --- | --- | --- | --- | --- | --- | --- |
|  | Indirect | n=52 | Direct | n=64 | Total | n=116 |  |
|  | Mean/Freq | %/SD | Mean/Freq | %/SD | Mean/Freq | %/SD |  |
| Age | 76.3 | 6.7 | 74.6 | 6.2 | 75.4 | 6.5 | 0.16 |
| BMI | 29.8 | 7.5 | 28.7 | 4.8 | 29.2 | 6.2 | 0.35 |
| Male | 36 | 69.2 | 40 | 62.5 | 76 | 65.5 | 0.73 |
| Race (White) | 46 | 88.5 | 63 | 98.4 | 109 | 94.0 | 0.69 |
| Diagnosis |  |  |  |  |  |  | 0.30 |
| ET | 43 | 82.7 | 50 | 78.1 | 93 | 80.2 |  |
| TDPD | 9 | 17.3 | 14 | 21.9 | 23 | 19.8 |  |
| Treatment Side (Left) | 41 | 78.8 | 46 | 71.9 | 87 | 75.0 | 0.39 |
| Preoperative SDR | 0.51 | 0.10 | 0.53 | 0.08 | 0.52 | 0.09 | 0.38 |
| Medical History |  |  |  |  |  |  |  |
| Current Anticoagulant/ Antiplatelet Therapy | 18 | 34.6 | 26 | 40.6 | 44 | 37.9 | 0.51 |
| Smoking History | 27 | 51.9 | 30 | 46.9 | 57 | 49.1 | 0.59 |
| Alcohol Use | 28 | 53.8 | 25 | 39.1 | 53 | 45.7 | 0.11 |
| Hypertension | 36 | 69.2 | 43 | 67.2 | 79 | 68.1 | 0.81 |
| Congestive Heart Failure | 7 | 13.5 | 6 | 9.4 | 13 | 11.2 | 0.49 |
| Diabetes | 16 | 30.8 | 20 | 31.3 | 36 | 31.0 | 0.96 |
| History of TIA/ Stroke | 7 | 13.5 | 9 | 14.1 | 16 | 13.8 | 0.93 |
| Peripheral Vascular Disease | 9 | 17.3 | 9 | 14.1 | 18 | 15.5 | 0.63 |
| COPD/ Pneumonia | 10 | 19.2 | 10 | 15.6 | 20 | 17.2 | 0.61 |
| Psychiatric Comorbidities | 20 | 38.5 | 30 | 46.9 | 50 | 43.1 | 0.36 |
| Hx of Neoplasm (except BCC) | 17 | 32.7 | 20 | 31.3 | 37 | 31.9 | 0.87 |
| Osteoporosis | 7 | 13.5 | 8 | 12.5 | 15 | 12.9 | 0.88 |

### Procedural Efficiency

Procedural efficiency metrics were markedly improved in the direct targeting group, with fewer total sonications (5·2 ± 1·6 vs. 6·7 ± 1·4; p < 0·0001; Figure 2A) and fewer targeting adjustments (2·3 ± 1·5 vs. 3·2 ± 1·7; p = 0·0069; Figure 2B). Direct targeting had lower total sonication time (72·0 ± 23·1 seconds vs. 91·6 ± 24·3 seconds; p < 0·0001; Figure 2C) and significantly reduced overall procedural duration (24·12 ± 12·01 minutes vs. 39·12 ± 15·43 minutes, excluding initial MR imaging; p < 0·0001; Figure 2D). The average total energy was 37,312 ± 17,013 J in the direct targeting cohort compared to 48,596 ± 21,228 J in the indirect targeting cohort (p = 0·0025; Figure 2E).

**Figure 2.**
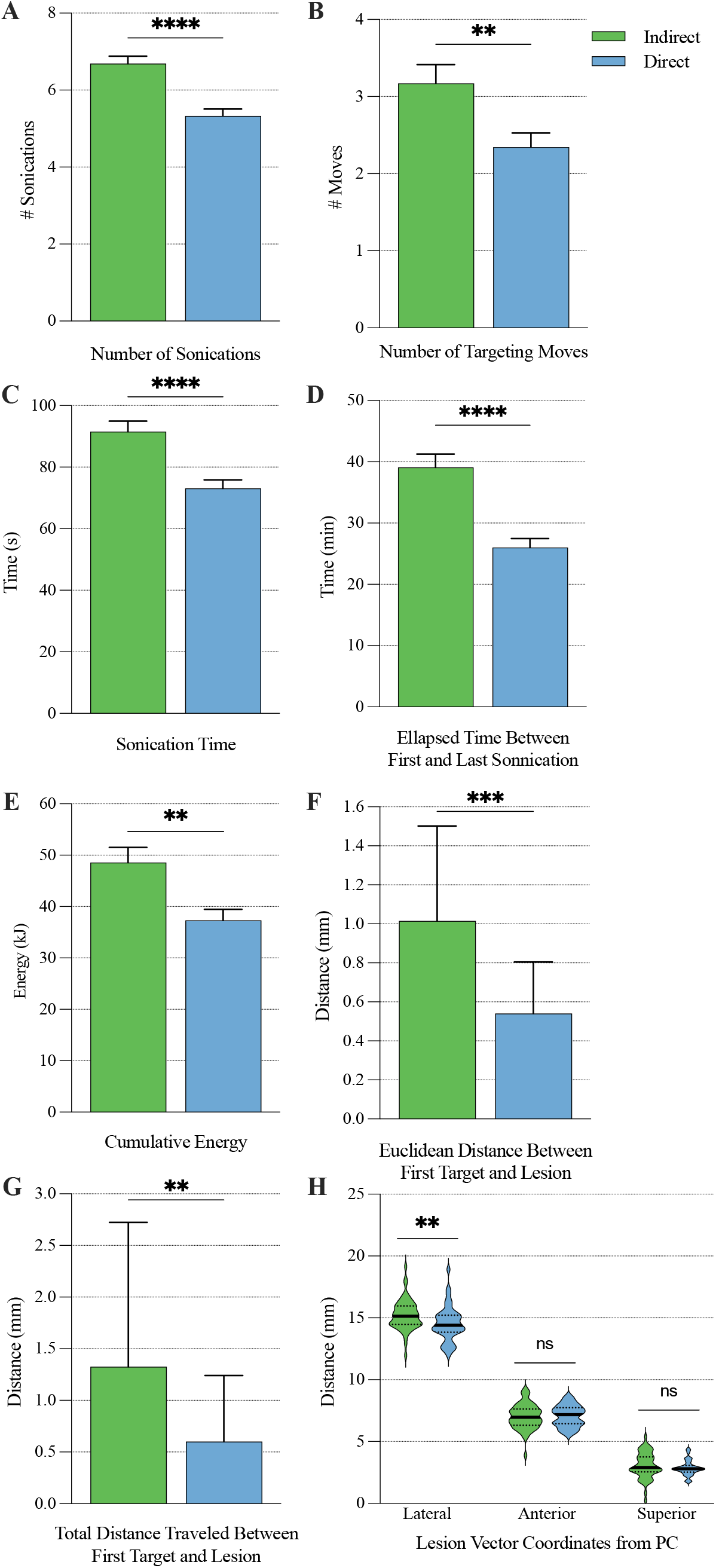
FGATIR direct targeting significantly improves MRgFUS procedural metrics compared to standard indirect targeting. Number of total sonications (A), number of targeting moves (B), total sonication time (C), elapsed time between the first and last sonication (procedural time, D), cumulative energy imparted into the parenchyma (E), the Euclidean distance between initial target coordinates and the coordinates of the first therapeutic dose sonication (F), and the total distance the target was moved between initial coordinates and the first therapeutic dose sonication (G) were all significantly reduced in the direct targeting cohort compared to the indirect targeting cohort. Lesion centers were more medial relative to the posterior commissure (PC) in the direct targeting cohort, but were comparable to the indirect cohort along the anteroposterior and superoinferior axes (H).

Direct targeting had reduced discrepancy between initial coordinates and the coordinates of the first therapeutic dose, with a median distance of 1·015 mm (IQR: 0·455-1·503 mm) and 0·540 mm (IQR 0·000-0·805 mm) for the indirect and direct targeting cohorts respectively (p = 0·0003; Figure 2F). The total distance of all targeting adjustments for direct targeting was 0·600 mm (IQR: 0·000-1·240 mm) compared to 1·325 mm (IQR: 0·585-2·723 mm) for indirect targeting (p = 0·0011; Figure 2G). The number of sonications was positively correlated with total cumulative energy imparted (Spearman’s ρ = 0·45, 95% CI: 0·2881–0·5901, p < 0·0001), size of the lesion (ρ = 0·2563, p = 0·0057), and extent of edema (ρ = 0·3583, p < 0·0001), with linear models indicating respective increases of∼8 mm^3^ lesion, ∼58 mm^3^ edema, and ∼66 mm^3^ total volume per additional sonication. However, sonication count alone was not correlated with amount of energy used for the permanent lesion (R^2^ = 0·011, p = 0·40).

### MRI-based Lesion and Edema Characteristics

Direct targeting was associated with significantly smaller lesion volumes (127·7 ± 47·6 mm^3^ vs. 162·0 ± 63·4 mm^3^; p = 0·0017; Figure 3A). Edema volumes were likewise reduced using direct targeting (902·0 ± 307·8 mm^3^ vs. 1260 ± 414·5 mm^3^; p < 0·0001; Figure 3B). Direct targeting lesions were centered 14·54 ± 1·43 mm lateral, 5·38 ± 1·39 mm posterior, and 2·87 ± 0·61 mm superior to the MCP. Lesion locations were slightly but consistently more medial than in the indirect targeting group without significant difference along anteroposterior or dorsoventral axes (Figure 2H).

**Figure 3.**
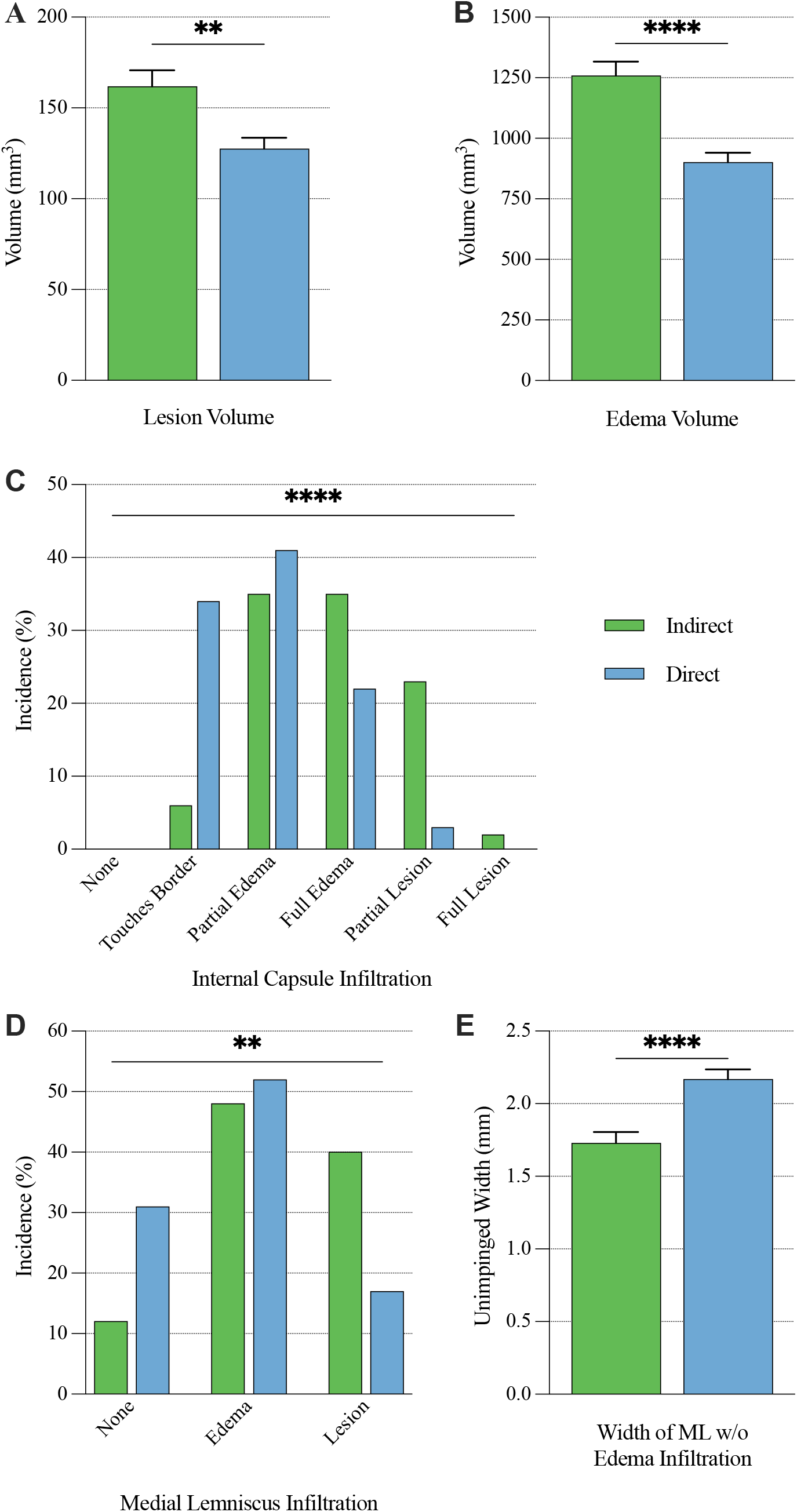
FGATIR direct targeting significantly improves lesion and perilesional edema volumetric metrics compared to standard indirect targeting. Both lesion (A) and perilesional edema (B) volumes were significantly reduced in the direct targeting cohort relative to the indirect targeting cohort. Impingement upon both the internal capsule (C) and the medial lemniscus (D) was correspondingly significantly reduced in the direct targeting cohort. The anteroposterior width of medial lemniscus that remained free of lesion or edema infiltration was significantly greater in the direct targeting cohort (E).

Direct targeting had less impingement on adjacent white matter tracts. Internal capsule involvement showed a significant shift toward lower scores and less IC impingement with direct targeting (p < 0·0001, Figure 3C). Similarly, edema impingement on the medial lemniscus was decreased with direct targeting (p = 0·0011; Figure 3D), and the width of ML was significantly greater with direct targeting (2·2 ± 0·5 mm vs. 1·7 ± 0·5 mm; p < 0·0001; Figure 3E). In contrast, the full width of the superolateral RW was nearly universally covered by lesion in both groups (p = 0·24).

### Tremor Outcomes

On POD1, both cohorts reported high tremor relief, with 93% of the direct group and 95% of the indirect group indicating significant tremor suppression (Figure 4A,B). These proportions had decreased to 80% and 85% by one month, and 72% and 59% by three months for the direct and indirect targeting cohorts respectively. No significant difference in reported tremor relief was found between either cohort at any time point (POD1: p = 0·14; one month: p = 0·70; three months: p = 0·40). However, quantitative spiral analysis demonstrated significantly greater tremor reduction for the direct targeting group (94·07 ± 8·93% vs. 79·65 ± 25·58%; p = 0·0099; Figure 4C).

**Figure 4.**
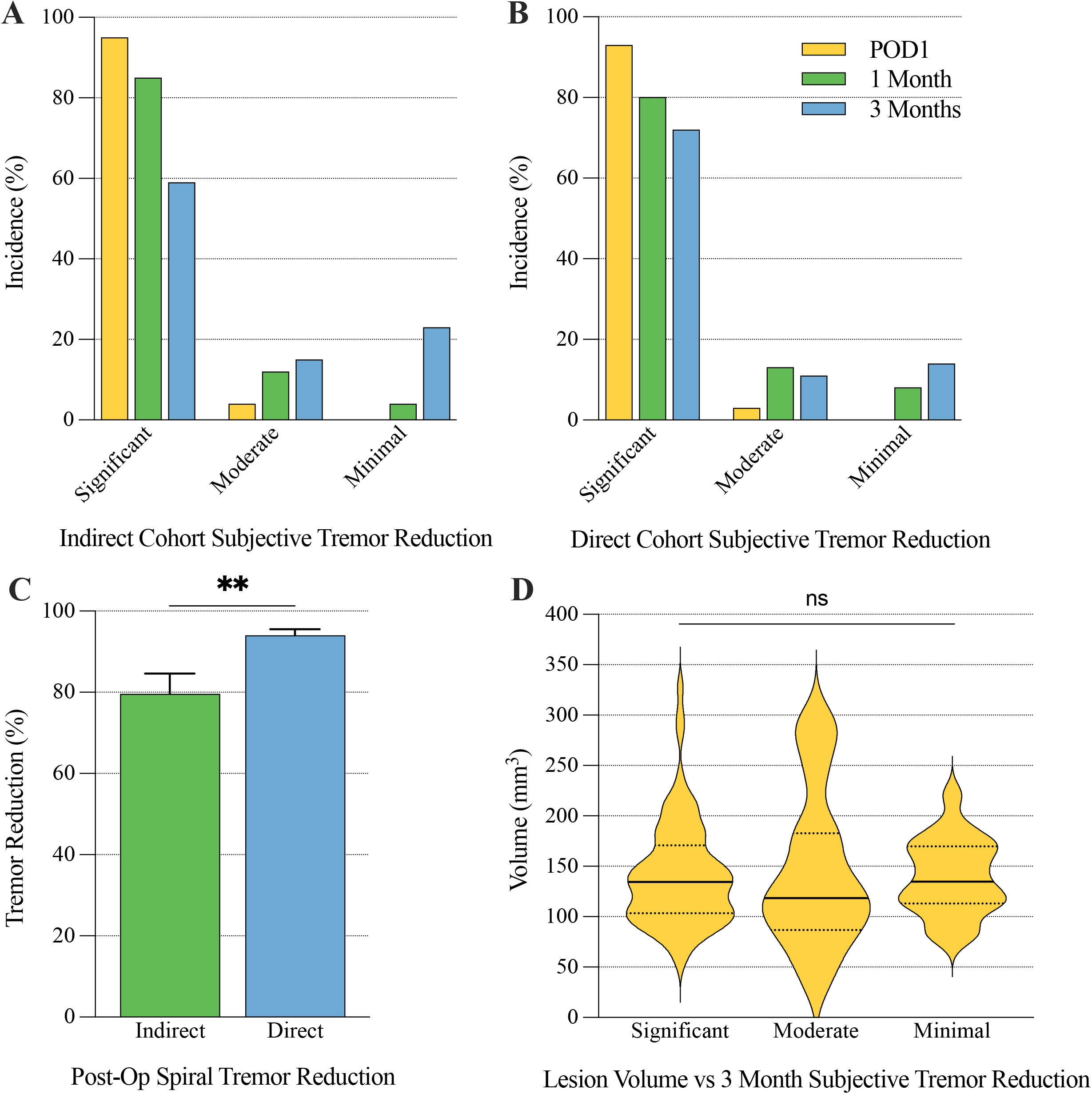
FGATIR direct targeting produces improved qualitative and equivalent subjective tremor outcomes compared to standard indirect targeting. Rates of patient reported significant, moderate, and minimal tremor reduction were not significantly different between the indirect (A) and direct (B) targeting cohorts at any timepoint. However, quantitative analysis of pre- and post- operative spiral drawings demonstrated significantly greater improvement in the direct targeting cohort (C). There was no correlation between postoperative day one lesion volume and three-month tremor outcomes (D).

### Incidence and Predictive Factors of Postoperative Neurological Adverse Effects

Neurologic adverse effects were most prevalent on POD1 and decreased over time. On day one, balance deficits were noted in 60% of patients in the direct targeting group and 80% in the indirect targeting group (p = 0·040). Strength deficits were present in 8% of the direct cohort and 27% of the indirect cohort (p = 0·010), and sensory deficits were reported in 32% of the direct cohort and 63% of the indirect cohort (p = 0·0010). Speech and swallowing difficulties were less frequent and proportionately not different between groups (Figure 5A). Balance remained a reported issue for a quarter of all patients at three months, while ongoing strength or sensory issues were each reported in roughly 10% of patients, speech deficits in 7%, and swallowing difficulties in 2% (no significant difference between groups; Figure 5B,C).

**Figure 5.**
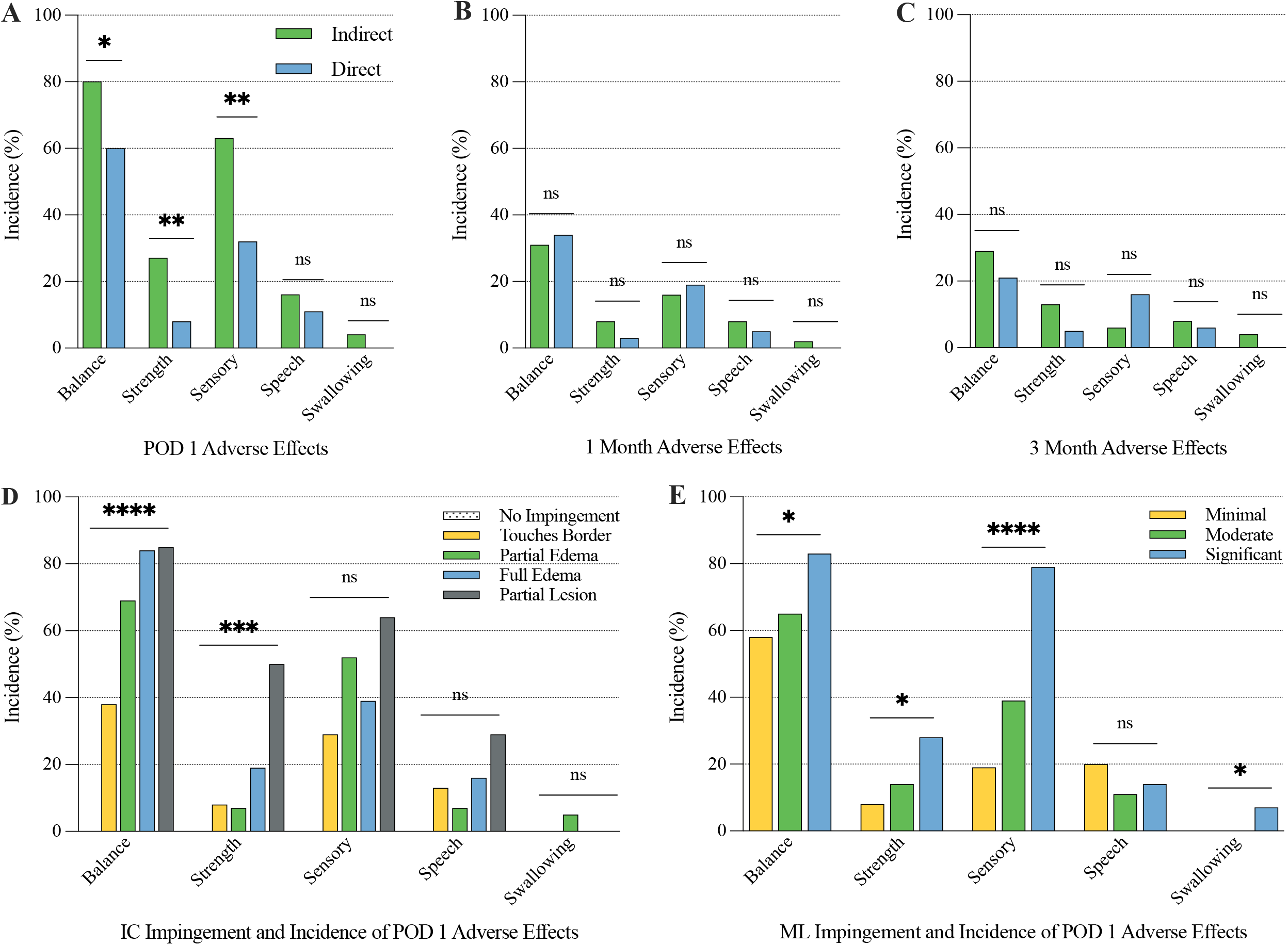
FGATIR direct targeting significantly reduces rates of acute neurological adverse effects compared to standard indirect targeting. The incidence of postoperative balance, strength, and sensory deficits were significantly lower for the direct targeting cohort, while the incidence of speech and swallowing deficits were comparable to those seen in the indirect cohort (A). There was no difference in the incidence of chronic adverse effects between the two cohorts at one-month (B) and three-month (C) assessments. Rates of balance and strength deficits were significantly correlated to degree of lesion and edema impingement of the internal capsule (D) and to a lesser extent the medial lemniscus (E), while rates of sensory and swallowing deficits were only correlated to impingement of the medial lemniscus (E).

Lesion volumes were not predictive of subjective tremor improvement at POD1 (p = 0·8740), one month (p = 0·65; Supplementary Figure 3B,C), or three months (p = 0·92; Figure 4B). Similarly, edema volumes were likewise not predictive of subjective tremor outcomes at any timepoint (POD1: p = 0·20; one month: p = 0·44; three months: p = 0·65; Supplementary Figure 3D-F). However, logistic regression analysis showed that lesion volume and edema volume were independent predictors of early postoperative adverse effects. At POD1, both volumes were strongly associated with the presence of any neurological adverse effect (lesion: OR = 1·231 per 10 mm^3^, 95% CI: 1·009–1·035, p = 0·0003; edema: OR = 1·221 per 100 mm^3^, 95% CI: 1·001–1·004, p = 0·0009). At three months, the POD1 lesion volume showed a significant association with persistent side effects (OR = 1·1046 per 10 mm^3^, 95% CI: 1·002–1·018, p = 0·0093), while POD1 edema volume was not a significant predictor of late side effects (p = 0·073).

A trend analysis was conducted to identify relationships between adjacent tract involvement and specific postoperative deficits. Increasing IC impingement scores were significantly associated with higher rates of postoperative balance deficits (p < 0·0001) and motor weakness (p = 0·0008), but were not significantly associated with sensory disturbances (p = 0·11), speech difficulty (p = 0·11), or swallowing impairment (p = 0·66; Figure 5D). ML involvement was predictive of multiple neurological outcomes, including balance impairment (p = 0·044), motor weakness (p = 0·043), and swallowing difficulty (p = 0·047); however, the strongest association was observed between ML involvement and sensory deficits (p < 0·0001), consistent with the known somatosensory role of this tract. ML involvement was not significantly associated with speech deficits (p = 0·58; Figure 5E). Further supporting the trend analysis, a greater width of ML spared from edema infiltration was significantly associated with lower odds of postoperative sensory deficits (OR = 0·290, 95% CI: 0·129–0·599, p = 0·0006), with moderate discriminative power (AUC = 0·697, Tjur R^3^ = 0·104), although the width of spared ML was not predictive of weakness (OR = 0·511, p = 0·14), balance (OR = 0·593, p = 0·15), speech (OR = 1·059, p = 0·90), or swallowing deficits (OR = 0·754, p = 0·82).

## Discussion

In this study, thalamotomy direct targeting enabled by FGATIR substantially improved the precision of lesion placement, enhancing both efficiency and safety of MRgFUS treatment of ET and TDPD tremors. Compared with conventional indirect targeting, the superior gray-white matter contrast of FGATIR yielded significantly smaller lesions (21% reduction) and perilesional edema (28% reduction), while still achieving superior tremor control on quantitative spiral analysis and equivalent patient-reported outcomes. This image-level precision was further accompanied by lower tract-specific impingement scores, preservation of ML width, and fewer acute adverse effects. In the same vein, FGATIR-based direct targeting significantly improved procedural efficiency by reducing the number of sonications, number of adjustments, cumulative energy imparted, and overall procedure time. Taken together, our study adds to the growing body of evidence that lesion localization is the primary determinant of tremor control, and that improved procedural efficiency enhances both safety and accessibility.

The relation between lesion volume and tremor control remains debated. Some studies associated larger lesions with superior tremor control,^21^ while others found no such association.^22,23^ Using quantitative analysis, our study found up to 14% greater tremor reduction in the FGATIR cohort despite smaller lesions, further supporting the primacy of precision over size. These findings align with Chua et al.^24^ who identified an inferoposterior Vim subregion “sweet spot” overlapping the Vim-DRTT junction as essential for tremor suppression. When both lesion size and location were jointly modeled, only the latter accounted for differences in outcomes.^24^ They further localized the average centroid of the junction to 14·75 ± 1·18 mm lateral, 5·82 ± 0·78 mm posterior, and 2·44 ± 0·57 mm superior to the MCP,^24^ which very closely aligns with our FGATIR targeted lesion centers, supporting the hypothesis that the thalamic-RW interface targeted using FGATIR represents the Vim-DRTT junction.

Volumetric reductions were not merely technical improvements but clinically meaningful. Our direct targeting lesions were 30-73% smaller compared to previous reports utilizing indirect targeting approaches^22,24^ and 6-48% smaller than those reported using other direct targeting modalities.^25,26^ Consistent with previous studies,^21-24^ lesion volume predicted procedure-related neurological morbidity across time points. Of note, edema correlated only with transient neurologic deficits in our study, which represents the first time, to our knowledge, its significant contribution to early clinical symptomatology has been empirically demonstrated. In keeping with what others have found,^24^ tract-specific correlations underscored the importance of precision: internal capsule involvement predicted weakness and imbalance while medial lemniscus involvement predicted sensory deficits. By minimizing tract impingement, FGATIR-based targeting can offer better safety profiles with reduced early neurological morbidity, particularly in a frail patient population.

Multiple studies have linked excessive sonication energy and prolonged procedures to greater edema, inferior outcomes, and higher complication rates.^11,21^ Direct targeting significantly streamlined our clinical workflow by reducing sonication number, total energy, and halving the iterative adjustments. Beyond limiting tissue disruption, enhanced procedural efficacy has implications both at patient- and systems-levels. Clinically, shorter procedures reduce discomfort and improve tolerability, especially for patients with musculoskeletal pain or claustrophobia. At the systems level, reducing procedural time can greatly enhance MRgFUS sustainability by reducing resource utilization, increasing throughput, and lowering per-case costs. Given the minimal barriers to implementation, widespread FGATIR adoption could enhance safety, reproducibility and access by standardizing procedural efficiency.^27,28^

Previous attempts at advancing targeting strategies highlight FGATIR’s pragmatic balance of precision, safety and scalability. Tractography-based methods, including the four-tract probabilistic approach, offer benefits in targeting reproducibility,^29,30^ but at the expense of higher sonication counts and longer procedures, often lasting 70 minutes or more. Furthermore, tractography approaches remain limited by processing time, diffusion signal quality, and variability in processing pipelines. Normative atlas-based approaches can identify lesional “sweet spots” but lack validated patient-specific adaptability.^23^

Other direct-targeting modalities such as fractional anisotropy T1 (FAT1) weighted and white-matter-nulled magnetization prepared rapid gradient echo (WMn-MPRAGE) imaging have shown feasibility in visualizing the Vim and DRTT,^25,26^ but these small single-center case series offer limited procedural reporting, lack rigorous outcomes validation, and require significant preprocedural processing. Ultra-high field 7T imaging and quantitative susceptibility mapping provide excellent anatomical delineation,^16,17^ but remain largely investigational and unavailable in most routine practices. Functional connectivity-based parcellations can accurately detect thalamic heterogeneity,^12,13,15^ but are derived from small healthy cohorts and show variable alignment with histological atlases. Against this backdrop, FGATIR emerges as a scalable strategy deployable on standard 1.5 - 3T scanners without additional software or processing burdens, that is outcome-validated in our retrospective cohort, and directly links anatomical precision to smaller lesions, fewer neurological deficits, and shorter procedures.

This study has limitations. It was a retrospective, single-center cohort treated by a single surgeon, which may restrict generalizability. The patient population markedly skewed older, white, and male. Follow-up was limited to three months, restricting conclusions about long-term durability or delayed adverse events. While patient-reported outcomes were supplemented with quantitative spiral analysis, standardized tremor rating scales such as CRST were not consistently available. Future multicenter, prospective studies are needed to confirm reproducibility across operators and institutions, and evaluate long-term outcomes.

In conclusion, we believe FGATIR direct targeting offers clear advantages over current targeting modalities. It is a clinically tested, widely deployable strategy with potential extension to other ablative and neuromodulatory interventions as neurosurgery advances toward individualized care.

## Data Availability

All data produced in the present study are available upon reasonable request to the authors

## Contributors

JC: study design, procedural assistance, study organization, medical record review, radiological assessment, spiral processing, data analysis, manuscript drafting, manuscript editing, figure creation. NU: medical record review, demographic analysis, manuscript drafting input, manuscript editing. AT: development of spiral processing script. HM: manuscript drafting input, manuscript editing. MH: study organization. AL: manuscript editing, procedural assistance. DC: study conceptualization, study design, manuscript drafting input, manuscript editing, procedural oversight. All authors had full access to all data included in the study, and had final responsibility for the decision to submit for publication. Both JC and NU accessed and verified the data.

## Declaration of Interest

We declare no competing interests.

## Data Sharing

Deidentified participant data, data dictionary, data analysis, and patient consent form will be made available with publication upon reasonable written request.

## Acknowledgements

The authors would like to thank the clinical and technical staff and students at Oregon Health & Science University for their support in conducting this work, as well as the patients and families who participated in this study. Specifically, we are grateful for the work of Suad Arnautovic, Christine Larsen, Molly Joyce, Beck Shafie, Haley Smith, Zoe Vanderhoek, and Tim Valuev.

---

**Supplementary Figure 1. Examples of radiological assessment.**Top) Orienting examples of postoperative day one lesions as visualized on FGATIR sequences. (Middle) Example measurements of perilesional edema volume (left and center) and medial lemniscus width without edema or lesion impingement (right) using FGATIR sequences. (Bottom) Examples of lesion volume measurements using T1 sequences.

**Supplementary Figure 2. Internal capsule and medial lemniscus impingement scoring rubrics.**(Top) Examples of the 6-point ordinal scoring for internal capsule impingement. There were no examples of no IC involvement (score 1) in the dataset; only scores 2-6 are shown. (Bottom) Examples of the 3-point ordinal scoring for medial lemniscus impingement.

**Supplementary Figure 3. There was no correlation between POD1 lesion or perilesional edema volumes and subjective tremor control at any timepoint.**Subjective tremor outcomes were found to have no correlation to either lesion (top row) or combined lesion and edema (bottom row) volumes at POD1 (A,B), one-month (B,C), or three-month (C,D) assessments.

## Notes

### Competing Interest Statement

The authors have declared no competing interest.

### Funding Statement

This study was funded by OHSU Parkinson Center Pilot Program and the Oregon Medical Research Foundation (#1029114).

### Author Declarations

The study protocol was reviewed and approved by Oregon Health & Science Institutional Review Board (STUDY00026905).

