## Supplementary figures and images for "Direct targeting for focused ultrasound thalamotomy in the treatment of movement disorders: a retrospective cohort study"

### Supplementary Figure 1

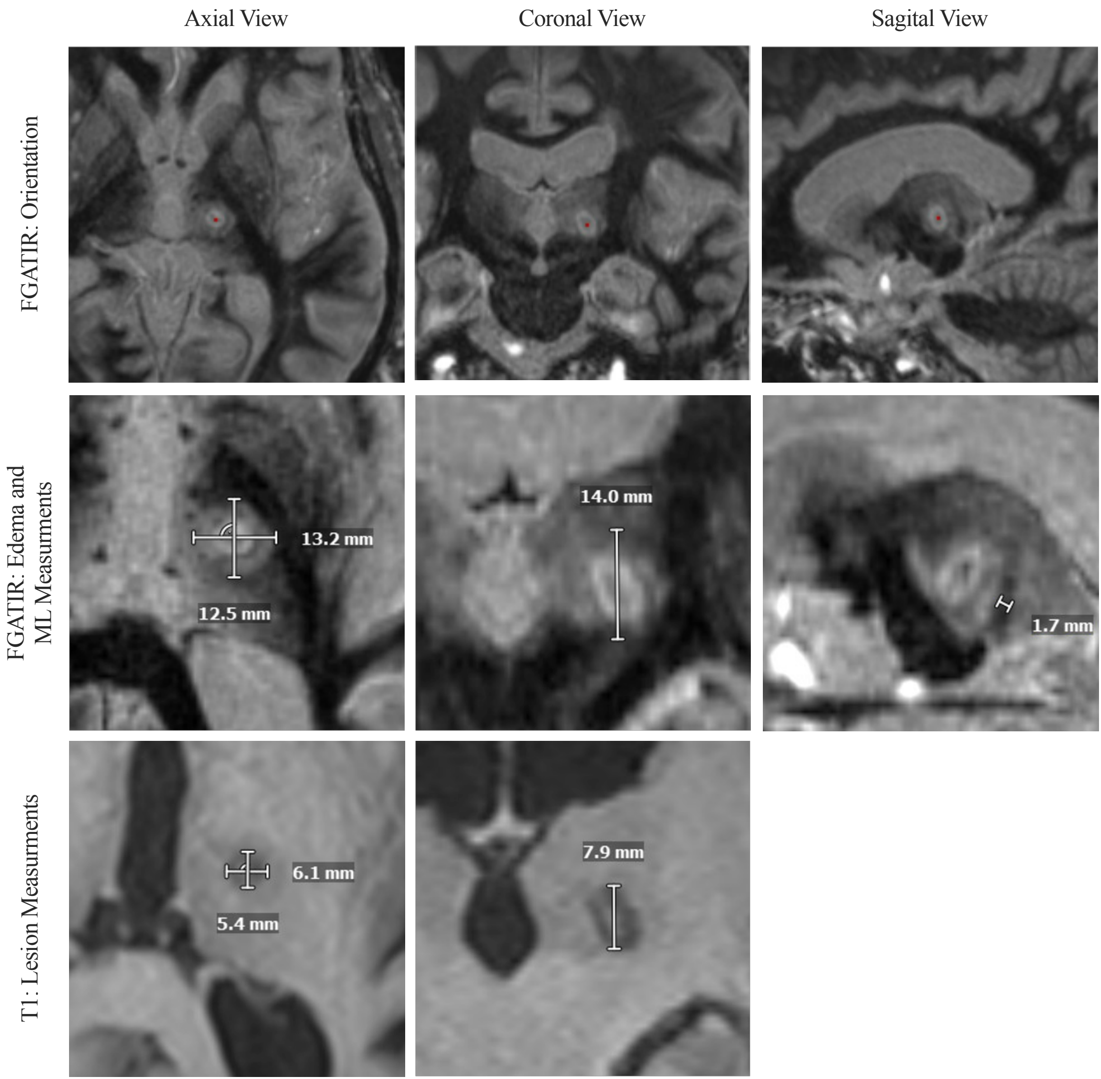

### Supplementary Figure 2

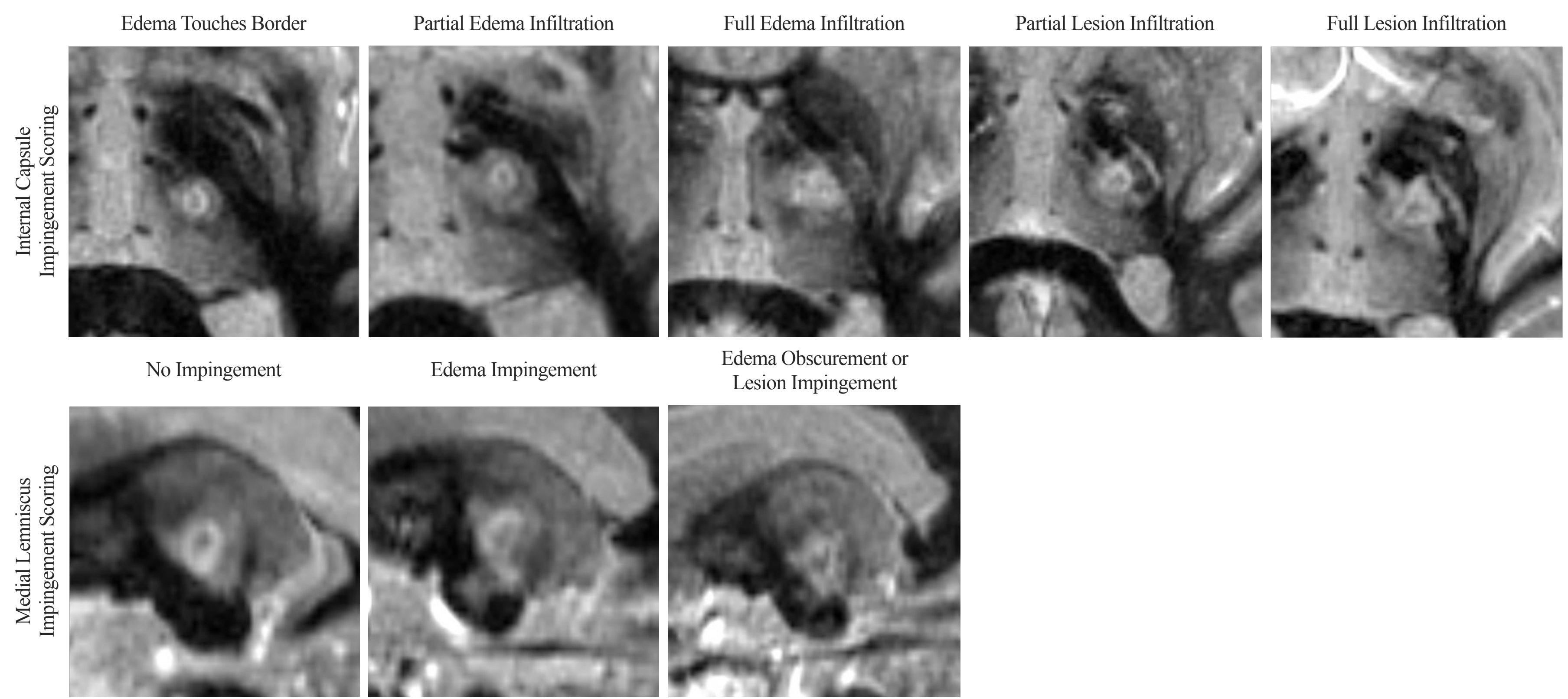

### Supplementary Figure 3

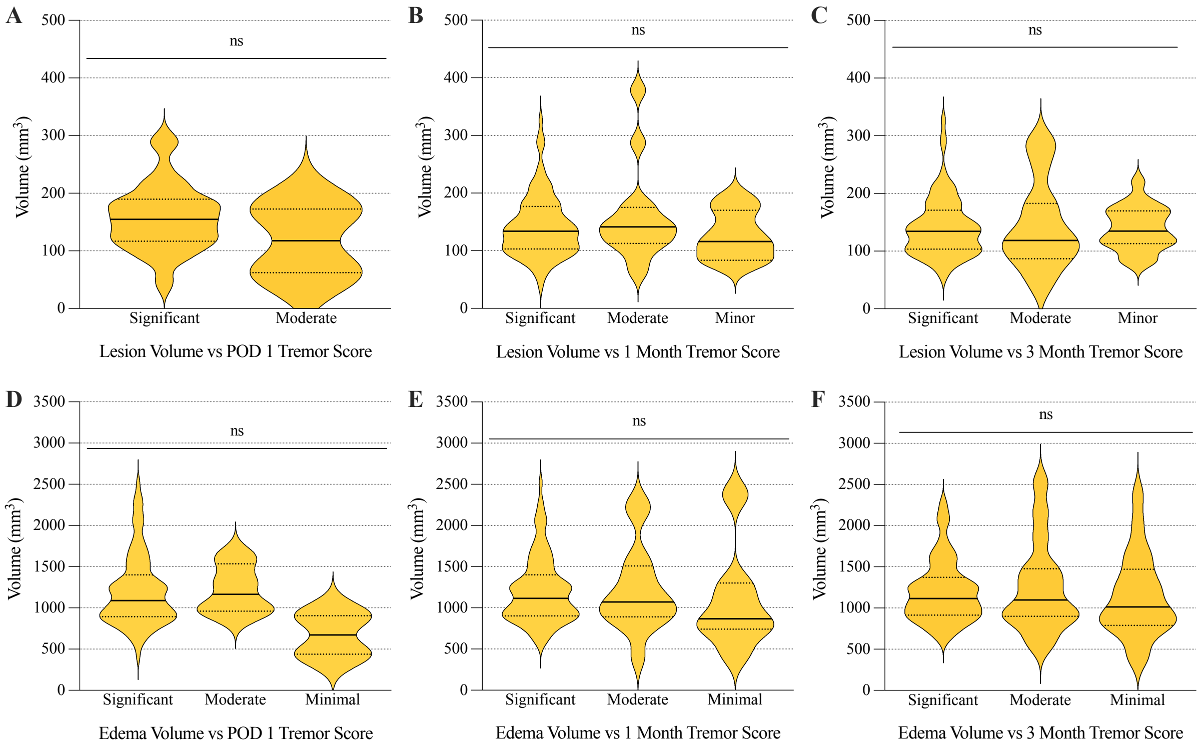
